# Determining the feasibility of randomising infants, children and young people to invasive and non-invasive urine sampling techniques

**DOI:** 10.64898/2026.08.22.26361091

**Authors:** Thomas Waterfield, Paula Taylor Miller, Clíona McDowell, Ashley Agus, Lynn Murphy, Charlotte Sanders, Anna Kearney, Frances Sherrett, Joy Wyche, Stuart Hartshorn, Srinivas Bandi, Bronagh Blackwood, Nefyn Williams, Damian Roland, Kathryn Ferris, Andrew Marshall, Mike Clarke, Alastair Sutcliffe, Kerry Woolfall

## Abstract

**Background:** Obtaining uncontaminated urine samples from children can be difficult. Clean catch urine (CCU) is non-invasive but may be slow and lead to a contaminated sample, whereas transurethral bladder catheterisation (TUBC) and suprapubic aspiration (SPA) are invasive. We assessed the feasibility of randomising children to a definitive trial.

**Methods:** FROG was a multicentre, randomised feasibility trial with a mixed-methods perspectives study, health-economic analysis and stakeholder consensus meeting. Children under 16 years requiring urine testing for suspected urinary tract infection (UTI) who could not provide a midstream sample were eligible for the feasibility trial. Parents, children and healthcare professionals were eligible for the perspectives study and consensus meeting.

**Results:** Of 703 children screened, 170 were offered the study and 99 were recruited. Overall, 64/170 (37.6%) consented to randomisation, exceeding the feasibility threshold (33%); 32 were allocated to CCU and 32 to TUBC. The allocated method was received by 46/64 (71.9%); delays, unsuccessful collection and distress contributed to non-receipt. Among participants with available cultures, contamination occurred in 2/12 (16.7%) allocated CCU and 0/6 allocated TUBC. No participants consented to randomisation involving SPA.

The perspectives study included 14 parent interviews, 89 parent questionnaires and 28 staff across 5 focus groups and 1 interview. CCU and TUBC were considered acceptable, although participants balanced speed and accuracy against pain and distress. SPA availability and acceptability were limited. A total of 19 stakeholders attended the consensus meeting; 94% supported recruiting children aged under 18 months and 100% supported comparing CCU with TUBC, without SPA. Accuracy was the highest-ranked outcome.

**Conclusions:** A definitive trial comparing CCU-first with TUBC-first in children aged under 18 months is feasible. Its primary outcomes should reflect diagnostic accuracy and clinical consequences of contamination, with successful collection, collection time, pain and distress assessed as key secondary outcomes.

**Plain English Summary:** Urinary tract infections are common in children, but accurate diagnosis depends on obtaining a reliable sample of urine. During collection, bacteria from the skin or surrounding area can get into the sample. This is called contamination and can make it appear that an infection is present when it is not. This mistake may lead to extra tests, delays in making treatment decisions and the unnecessary use of antibiotics.

Internationally, three methods are used to collect urine from young children, and we do not know which method is best. A “clean catch” sample involves catching urine in a sterile container. It does not hurt, but collection can be slow and contamination of the sample is not unusual. A catheter sample uses a tube passed into the child’s bladder, while a third method uses a needle passed into the bladder through the skin. Catheter and needle sampling may provide cleaner samples more quickly but can cause pain or distress.

We examined whether families would join a study comparing these three methods and whether a larger trial would be practical and acceptable. We screened 703 children and offered the study to 170 families. Overall, 99 families took part and 64 agreed that their child could be randomly allocated to clean catch or catheter sampling, which was more than we expected. No family agreed to randomisation involving needle sampling. A total of 46 of the 64 (71.9%) children received their allocated method. Delay, unsuccessful collection and distress contributed to others receiving a different method. Among children with available laboratory results, 2 of 12 clean catch cultures were contaminated compared with none of 6 catheter cultures.

We spoke with 14 parents and 28 healthcare professionals, and collected data from 89 parent questionnaires to gather their views about the study. They told us that both clean catch and catheter sampling were acceptable, but participants balanced accuracy (obtaining a reliable sample) and speed against the pain and distress. A meeting of 19 parents and professionals supported the proposal to do a larger study to compare clean catch with catheter sampling in children aged under 18 months attending emergency departments. Accuracy was considered the most important outcome.

This shows that a larger trial is feasible. It should assess diagnostic accuracy and whether contamination leads to further healthcare contacts, repeat sampling or antibiotics. Successful collection, collection time, pain and distress should also be measured.

## Introduction

Urinary tract infections (UTIs) are the second most common serious bacterial infection in children and account for a large number of presentations to primary and secondary care [1]. By 16 years of age, approximately 1 in 10 girls and 1 in 30 boys will have experienced a UTI [2]. Clinical features are often non-specific, particularly in younger children, and include fever, vomiting, abdominal pain and lethargy [3,4]. Urine testing is required to guide antibiotic treatment and follow-up, and prompt diagnosis and treatment are important to reduce complications including sepsis and renal scarring [5].

The National Institute for Health and Care Excellence (NICE) recommends clean catch urine (CCU) collection as the first line approach [3]. Invasive methods, including transurethral bladder catheterisation (TUBC) and suprapubic aspiration (SPA), are recommended only when non-invasive methods are not possible or practical [3]. Non-invasive sampling is painless, acceptable to families and feasible in primary care, but can be time consuming and is associated with high rates of bacterial contamination. Contamination can lead to false positive results, unnecessary antibiotics, further investigations, avoidable hospital attendance or admission, and poorer antimicrobial stewardship.

Existing evidence suggests that contamination is higher with non-invasive than invasive sampling. Reported contamination rates range from 26% to 36% for CCU, compared with approximately 12% for TUBC and 1% for SPA [10–13]. International practice varies, with many European and North American guidelines favouring invasive sampling in younger children because of the lower risk of contamination [4,6–9].

This variety of practice is important in the context of antimicrobial resistance. The World Health Organization has identified antimicrobial resistance as one of the greatest threats facing humanity [14]. Childhood UTIs are most commonly caused by Escherichia coli, and resistance is increasing because of over-use of antibiotics. In England, approximately 30% of E. coli urinary isolates are resistant to trimethoprim and 10% to cefalexin [3,15]. Reducing unnecessary antibiotic use is therefore a priority. False positive urine culture results may expose children to unnecessary treatment and follow-up; in infants under three months, suspected UTI can also lead to septic screening, lumbar puncture, admission and parenteral antibiotics [3].

A definitive randomised trial is needed to compare reliably the effects invasive and non-invasive urine collection methods. However, before such a trial can be undertaken, it is necessary to establish whether such it would be feasible and acceptable. We conducted the FROG feasibility study to assess recruitment, randomisation, intervention delivery and acceptability of CCU, TUBC and SPA, and to inform the design, population, outcomes and resource requirements of a future definitive trial.

## Methods

The FROG study was a pragmatic, multicentre, randomised feasibility trial with an embedded health economic component, a mixed-methods perspectives study and a stakeholder consensus meeting to inform the design of a definitive trial. We published the full study design prospectively [16].

### Feasibility trial design

We conducted a pragmatic, multicentre, randomised feasibility trial and report it in accordance with the CONSORT 2010 extension for randomised pilot and feasibility trials [17].

#### Recruitment and sampling

We recruited trial participants from six UK hospitals in clinical areas where children commonly undergo urine testing for suspected UTI, including paediatric emergency departments, paediatric assessment units, inpatient wards and outpatient clinics. We did not recruit from neonatal units.

Children were eligible if they were aged under 16 years, required urine testing for suspected UTI and were unable to provide a midstream urine sample because they were not toilet trained. We excluded children if they required immediate invasive sampling; both invasive methods (TUBC and SPA) were inappropriate or unavailable for them; they were sedated or admitted to intensive care; language barriers could not be overcome; no parent or legal representative was available; or consent was declined.

Clinical teams identified potentially eligible children during routine care. Following screening procedures, a clinical team member introduced the study to eligible children and their parents. The research team provided interested families with verbal and written information, along with a pre-recorded information video. We provided children with age-appropriate information where appropriate. Parents or legal representatives provided written informed consent before randomisation, and we sought assent from children where appropriate. Families who declined to take part received usual care and, where they agreed, we recorded their reasons for non-participation.

#### Interventions

We offered participants allocation to CCU, TUBC or SPA (where available). CCU involved non-invasive urine collection in a sterile container. TUBC involved passage of a flexible catheter through the urethra into the bladder. SPA involved needle aspiration of urine through the lower abdominal wall. Clinical staff performed the procedures according to local policy and usual practice. The child or parent could request that clinical staff discontinue, modify or replace sampling with an alternative method; clinical staff could also do so according to clinical judgement. We recorded adherence, discontinuation and crossover.

#### Outcomes

We defined the primary feasibility outcome as the proportion of participants offered the study who consented to randomisation. We set the pre-specified feasibility threshold as consent to randomisation in more than one-third of those offered the study.

Secondary outcomes included the proportion of participants whom clinicians judged unsuitable for the study; consent to all three interventions or restricted two-arm randomisation; receipt of the allocated intervention; contamination by urine collection method; adverse events; time to urine sample collection; pain and distress; final diagnosis of UTI; and resource use and costs, including parent-reported resource use collected using an adapted Modular Resource-use Measure (ModRUM) questionnaire.

We applied a single microbiological definition to urine culture outcomes. We defined UTI as pure growth of a single organism at ≥10D CFU/mL with pyuria. We classified cultures with mixed growth at ≥10D CFU/mL, as contaminated. We classified cultures with growth below 10D CFU/mL as negative/no growth.

We assessed pain and distress after urine sampling using age-appropriate and parent-reported measures, including the Wong-Baker FACES Pain Rating Scale, the FLACC scale and the Subjective Units of Distress Scale, for all of which higher scores indicate worse pain or distress.

#### Sample size

As this was a feasibility study, we did not undertake a formal hypothesis-testing sample size calculation. We selected the planned sample size of 100 to provide sufficient information on screening, recruitment, consent, randomisation, intervention delivery and outcome collection to inform a future definitive trial.

#### Randomisation and allocation concealment

We randomised consenting participants using a secure automated web-based randomisation system. We allocated participants in a 1:1:1 ratio to CCU, TUBC or SPA. If one invasive method was contraindicated, inappropriate or unavailable, we randomised participants in a 1:1 ratio between CCU and the remaining available invasive method. We used randomly permuted blocks and concealed allocation until randomisation. We could not use blinding because of the nature of the interventions.

#### Statistical methods

We conducted descriptive analyses. We summarised screening, eligibility, consent, randomisation and retention using frequencies and proportions. Baseline characteristics were summarised by allocated group using medians and interquartile ranges, means and standard deviations, or frequencies and percentages, as appropriate. We did not plan formal between-group hypothesis testing and reported outcome-specific denominators where data were incomplete.

For the prespecified sensitivity analysis, participants were classified as higher risk if they had one or more of the following: age <3 months; fever >39°C within the preceding 24 hours; temperature >38°C at presentation; clinical signs of sepsis or septic shock; heart rate >150 beats/minute; capillary refill time >2 seconds; or previous UTI. We summarised randomisation by higher- and lower-risk classification.

#### Health economic analysis

We embedded an exploratory health-economic analysis from a hospital perspective within FROG to estimate costs and inform a future definitive economic evaluation. The analysis included all recruited participants with a recorded urine-collection method, including those who declined randomisation, grouped according to the method attempted. As no participant underwent SPA, we summarised resource use and costs for CCU and TUBC. We collected hospital resource use from case report forms and clinical records and obtained information on equipment, staff roles and procedure times from participating sites. We also piloted an adapted parent/guardian Modular Resource-Use Measure [18]. We applied UK unit costs and summarised mean cumulative costs at 7 days, 30 days and 6 months. Analyses were descriptive and followed CHEERS 2022 reporting guidance [19].

### Embedded mixed-methods perspectives study

An embedded mixed-methods perspectives study involving questionnaires, interviews and focus groups explored acceptability and feasibility of the trial design, urine collection methods, recruitment procedures and outcomes for a future definitive trial. Participants included parents or guardians and healthcare practitioners. KW (female, social scientist, PhD) used previous research and patient and public involvement feedback to develop information materials, questionnaires and topic guides which were developed iteratively [20,21].

#### Recruitment, sampling and conduct

Participants were recruited from feasibility trial sites, via social media and relevant UK-based organisations to support diversity in geography, ethnicity, age and previous urine sampling experience.

At participating sites, parents or guardians of children aged under 16 years, and children aged 7 to under 16 years, were eligible if they had been approached for the feasibility trial, including those who declined randomisation. After the trial recruitment discussion, practitioners invited parents and children to complete a brief questionnaire and to register interest in taking part in an interview at a later date (e.g. within a month). Translated information sheets and interpreters were available for interviews.

For recruitment through social media, email and relevant organisations, parents or guardians and children were eligible if they had experienced hospital urine testing for suspected UTI within the preceding three years. CS and JW distributed an advert containing details of the study and instructions on how to register interest in an online or face-to-face interview; face-to-face interviews were available to those living in North West England.

Once a parent and/or child had registered interest the researcher (JW, female, psychologist, MA or CS, female, social scientist, PhD) sent an age and language-appropriate Participant Information Sheet, checked eligibility through and online screening survey and whether an interpreter was required. Interviews could be individual or joint parent-child interviews, depending on preference and child assent.

Healthcare practitioners, including doctors, nurses, research staff and allied health professionals, were eligible whether or not they had been directly involved in the feasibility trial. CS, KW and PTM (female, chartered psychologist, PhD) sent invitations via email and a social media advert via professional networks and invited them to attend an online focus group. Individual interviews were offered if a practitioner could not attend a focus group

Once eligibility was checked, CS or JW sent parents and staff a list of outcomes and link to an online consent form to seek informed consent or child assent prior to interview or focus group participation. A paper version was available on request (e.g. to seek child assent).

CS, JW or KW used an age/ group appropriate topic guide to explore feasibility and acceptability of the trial, barriers and facilitators to recruitment, acceptability of invasive and non-invasive urine collection, eligibility criteria and important outcomes. Clinical vignettes were used to explore views across different patient groups and clinical scenarios.

Purposive sampling was used to include participants with different experiences, including those who consented to or declined randomisation, different urine collection methods, age groups, different sites and settings (e.g. staff from emergency and outpatient settings).

Based on previous studies we anticipated interviewing 25-35 participants (∼15-20 parents and ∼10-15 children) and 6-8 staff in one of up to five focus groups plus up to 10 interviews with staff unable to attend a focus group. We sampled to reach the point of information power, which considers factors including the study aims, sample specificity, use of theory and the quality of dialogue [22]. The Consolidated Criteria for Reporting Qualitative Research checklist was used to aid reporting.

#### Analysis

Interviews and focus groups were audio-recorded with consent, transcribed verbatim by a transcription company (UK Transcription, Brighton, UK), checked and anonymised. Qualitative data were managed using NVivo V.15 software for organising and coding and analysed using reflexive thematic analysis, informed by constant comparison [23]. KW, CS, FS and AK met regularly to develop the coding framework. Questionnaire data were inputted into SPSS version 28 and summarised descriptively. Findings were synthesised and mapped to the Adapted Framework of Acceptability [24].

### Consensus meeting

We held an online consensus meeting on 19 June 2026. We purposively recruited parents/carers, clinicians, research staff and methodological experts from the groups described in Figure 1, alongside co-investigators and subject experts. PTM and TW sent invitations via email to stakeholders to attend the consensus meeting. Parents of children who had taken part in the feasibility trial and registered their interest in being contacted, were also invited by telephone with verbal explanation of the meeting. Stakeholders were provided with a participant information sheet, preparation sheet for virtual consensus meetings and an opportunity to ask questions prior to providing informed consent. TW, KW and PTM facilitated the meeting.

**Figure 1.**
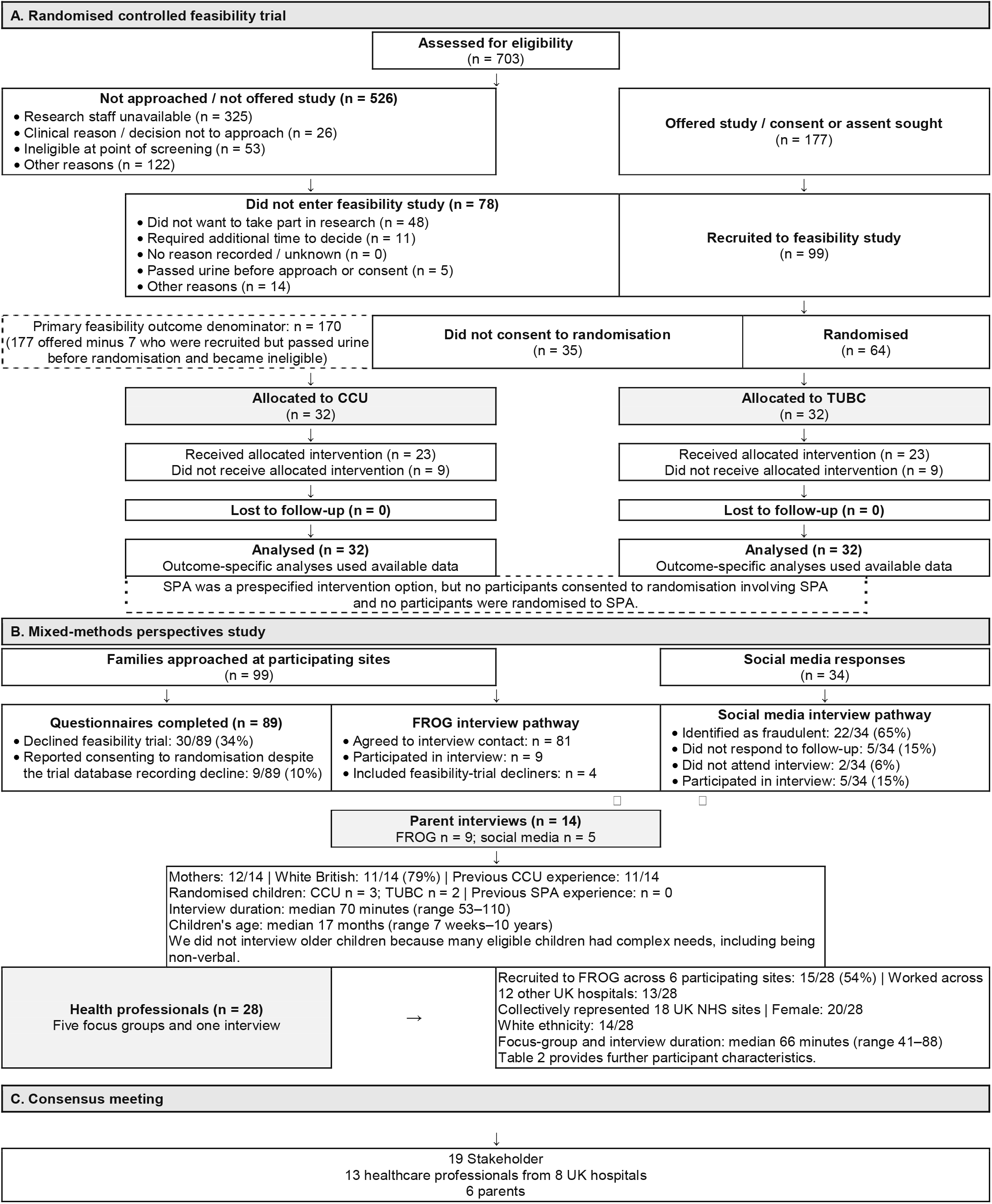
Participant flow through the FROG feasibility trial and mixed-methods perspectives study.

Following presentation of the feasibility trial and results and findings from the perspectives study, participants used structured discussion to develop the population, intervention, comparator and outcomes (PICO) for a definitive trial. Proposed statements on the population and intervention/comparator were refined through discussion and voted on anonymously using Slido. Consensus was defined prospectively as ≥70% agreement.

Potential outcome measures identified in the perspectives study were presented and ranked by participants.

### Patient and public involvement

Patient and public involvement contributors informed the study design, participant-facing materials, trial identity, outcome selection, conduct, interpretation and dissemination planning. Before submitting the grant application, we involved children, young people and parents through virtual meetings and surveys.

### Ethics and trial registration

The North East–Newcastle and North Tyneside 1 Research Ethics Committee approved the study. We prospectively registered the study with the International Standard Randomised Controlled Trial Number registry (ISRCTN84676764) on 1 April 2025.

## Results

### Participant flow

Participant flow is summarised in Figure 1. Recruitment took place between 8 June 2025 and 27 October 2025. Of 703 children screened, 170 were offered the study and 99 were recruited. A total of 64 consented to randomisation (32 CCU; 32 TUBC). No participants consented to SPA, and there were no losses to follow-up.

Of the 99 families approached at participating sites, 89 completed questionnaires and 9 participated in online interviews, including 4 who had declined randomisation. Among questionnaire respondents, 30/89 (34%) had declined randomisation, while 9/89 (10%) reported consenting although the trial database recorded that they had declined. Social media recruitment resulted in 5 additional parent interviews from 34 responses; 22/34 (65%) were fraudulent, 5/34 (15%) did not respond and 2/34 (6%) did not attend. Overall, 14 parents were interviewed. We did not interview children because potentially eligible children had complex needs, including being non-verbal.

A total of 28 health professionals participated in 5 online focus groups and 1 interview, collectively representing 18 UK NHS sites. Of these, 15 had recruited to FROG across its 6 sites, while 13 worked across 12 other hospitals. Figure 1 and Tables 1 and 2 provide further participant characteristics. Median duration of parent interviews was 70 minutes (53-110 minutes) and 66 minutes (41-88 minutes) for health professional focus groups and interviews.

**Table 1.** Baseline characteristics and outcomes by randomised allocation.

| CHARACTERISTIC OR OUTCOME | CCU (N=32) | TUBC (N=32) |
| --- | --- | --- |
| **BASELINE CHARACTERISTICS** |  |  |
| **FEMALE SEX** | 19 (59.4%) | 19 (59.4%) |
| **AGE, DAYS, MEDIAN (IQR)** | 262 (61–537) | 277 (50–624) |
| **ETHNICITY** |  |  |
| **WHITE** | 23 (71.9%) | 20 (62.5%) |
| **MIXED OR MULTIPLE ETHNIC GROUPS** | 0 (0.0%) | 5 (15.6%) |
| ASIAN OR ASIAN BRITISH | 8 (25.0%) | 5 (15.6%) |
| BLACK, BLACK BRITISH, CARIBBEAN OR AFRICAN | 1 (3.1%) | 0 (0.0%) |
| OTHER ETHNIC GROUP | 0 (0.0%) | 2 (6.3%) |
| RECRUITED FROM EMERGENCY DEPARTMENT | 30 (93.8%) | 31 (96.9%) |
| FEVER IN PRECEDING 24 HOURS | 21 (65.6%) | 20 (62.5%) |
| SIGNS OF SEPSIS | 5 (15.6%) | 5 (15.6%) |
| FEASIBILITY OUTCOMES |  |  |
| RECEIVED ALLOCATED URINE-COLLECTION METHOD | 23 (71.9%) | 23 (71.9%) |
| TIME TO URINE SAMPLE, MINUTES, MEDIAN (IQR) | 78 (39–182);<br>n=28 | 41 (25–68);<br>n=30 |
| CLINICAL OUTCOMES |  |  |
| CONTAMINATED URINE CULTURE | 2/12 (16.7%) | 0/6 (0.0%) |
| UTI BY MICROBIOLOGICAL DEFINITION | 4/12 (33.3%) | 3/6 (50.0%) |
| FINAL CLINICAL DIAGNOSIS OF UTI | 6 (18.8%) | 7 (21.9%) |
| FLACC PAIN SCORE, MEAN (SD) | 2.42 (3.01); n=26 | 6.17 (3.21);<br>n=30 |
| SUDS DISTRESS SCORE, MEAN (SD) | 2.20 (2.84); n=25 | 5.43 (3.14);<br>n=28 |
| ADVERSE EVENTS | 0 (0.0%) | 0 (0.0%) |

Data are n (%) unless otherwise stated. Outcome-specific denominators are shown where data were incomplete. CCU: clean catch urine; TUBC: transurethral bladder catheterisation; UTI: urinary tract infection; IQR: interquartile range; SD: standard deviation; FLACC: Face, Legs, Activity, Cry, Consolability; SUDS: Subjective Units of Distress Scale.

**Table 2.** Characteristics of perspectives-study participants.

| **Parents (N=14)** | **n (%)** |
| --- | --- |
| *Mother* | **12 (86)** |
| *Father* | **2 (14)** |
| *Asian or Asian British* | **3 (21)** |
| *White* | **11 (79)** |
| *Child age, median (range)* | **17 months (1.6–120)** |
| *Recruited from FROG trial site* | **9 (64)** |
| *Recruited from Social media* | **5 (36)** |
| *Randomised* | **5 (56)** |
| *Declined randomisation* | **4 (44)** |
| *Allocated CCU* | **3 (60)** |
| *Allocated TUBC* | **2 (40)** |
| **Healthcare professionals (N=28)** | **n (%)** |
| *Asian or Asian British* | **4 (14)** |
| *Black, Black British, Caribbean or African* | **1 (4)** |
| *White* | **14 (50)** |
| *Other ethnic group* | **2 (7)** |
| *Ethnicity not available* | **7 (25)** |
| *Female* | **20 (71)** |
| *Male* | **7 (25)** |
| *Gender not available* | **1 (4)** |
| *Recruited from FROG trial site* | **15 (54)** |
| *Recruited from other UK hospital* | **13 (46)** |
| *Role - paediatrician* | **12 (43)** |
| *Role - advanced nurse practitioner* | **3 (11)** |
| *Role - nurse* | **3 (11)** |
| *Role - research nurse* | **10 (36)** |

Data are n (%) unless otherwise stated. CCU: clean catch urine; TUBC: transurethral bladder catheterisation.

### Feasibility trial outcomes

The primary feasibility outcome was achieved: 64/170 (37.6%) children offered the study and included in the primary-outcome analysis consented to randomisation, exceeding the prespecified one-third threshold. The mean recruitment rate was 3.7 participants per site per month, exceeding the target of 3. Among randomised participants, median age was 275 days (IQR 61–546), 38/64 (59.4%) were female, 61/64 (95.3%) were recruited from an emergency department. Of the 99 recruited participants, 73/99 (73.7%) were classified as higher risk and 26/99 (26.3%) as lower risk. Randomisation was achieved in 49/73 (67.1%) higher-risk participants and 15/26 (57.7%) lower-risk participants. Table 1 presents ethnicity and other clinical characteristics.

Overall, 46/64 (71.9%) participants received their allocated urine-collection method: 23/32 (71.9%) in each group. Among those who did not receive CCU, the most common reason was that collection was taking too long (4/9). Reasons for not receiving TUBC included failure to obtain urine (3/9), passing urine before the procedure (3/9) and distress (2/9). The remaining cases reflected other logistical or undocumented reasons (n=6).

Urine samples were obtained from 28/32 participants allocated to CCU and 30/32 allocated to TUBC. All obtained samples underwent initial testing by dipstick analysis or microscopy. In accordance with routine clinical practice, only a subset was sent for culture, guided by the point-of-care or microscopy findings. Among participants whose samples were cultured, contamination was detected in 2/12 (16.7%) allocated to CCU and 0/6 allocated to TUBC. Median collection time was 78 minutes (IQR 39–182; n=28) with CCU and 41 minutes (IQR 25–68; n=30) with TUBC. Mean FLACC pain scores were 2.42 (SD 3.01; n=26) and 6.17 (SD 3.21; n=30), respectively.

Corresponding mean SUDS distress scores were 2.20 (SD 2.84; n=25) and 5.43 (SD 3.14; n=28). One participant allocated to TUBC completed the Wong–Baker FACES Pain Rating Scale; all other participants were either too young to self-report or unable to do so because of additional needs.

A final clinical diagnosis of UTI was recorded in 6/32 (18.8%) participants allocated to CCU and 7/32 (21.9%) allocated to TUBC. Microbiologically defined UTI was identified in 4/12 (33.3%) and 3/6 (50.0%) participants whose samples were cultured, respectively. No adverse events were reported.

### Health-economic analysis

The health-economic analysis included all recruited participants with a recorded urine-collection method, including those who declined randomisation, and grouped them by the method attempted. Data were available for 91/99 (91.9%) participants: 56 underwent or attempted CCU and 35 underwent or attempted TUBC. Only 16 ModRUM questionnaires were returned, which was insufficient for analysis.

During follow-up, additional urine sampling occurred in 9/56 (16.1%) participants in the CCU group and 6/35 (17.1%) in the TUBC group. Imaging was undertaken in 5/56 (8.9%) and 5/35 (14.3%), respectively, while antibiotics for UTI were prescribed to 11/56 (19.6%) and 8/35 (22.9%). Emergency department re-attendance occurred in 6/56 (10.7%) and 7/35 (20.0%), and hospital readmission in 1/56 (1.8%) and 3/35 (8.6%), respectively.

Mean cumulative hospital costs were £590.43 (95% CI 381.51-799.35) with CCU and £642.10 (427.11-857.09) with TUBC at 7 days, £680.37 (393.69-967.05) and £730.31 (481.64-978.98) at 30 days, and £751.11 (95% CI 452.20-1050.02) and £901.10 (95% CI 542.74-1259.45) at 6 months, respectively. By 30 days, 91% of 6-month costs in the CCU group and 81% in the TUBC group had accrued.

### Perspectives study

#### Acceptability of participation and randomisation

Parents participated for several reasons, including helping their child or other children, valuing research and trusting the clinician introducing the study:

> *“To her directly, I’m not really sure of any benefits, I suppose, but for me, having children and knowing the importance of research studies, then I would say, yeah, there’s a wider positive outcome for other children and going forward.”* (P45, mother, interview)

Some also hoped that TUBC might *“speed up the process”* (P14, mother, interview) or improve the likelihood of obtaining a sample following previous difficulties with CCU. Parents were generally approached within the first few hours of arrival. Some had already attempted CCU, while others were approached immediately.

Nevertheless, most parents recruited to the feasibility trial did not feel *“any kind of pressure”* (P33, mother, interview) to participate and had sufficient time to decide, even within 1 hour while their child was unwell. Although a minority found the decision difficult (Table 3, statement I), most described it as quick, easy and “*he was going to be tested anyway* “(P54, mother, interview)

**Table 3.** Parental consent decision making in the FROG Study n=89.

| Statement |  | Agree | Neither agree nor disagree | Disagree |
| --- | --- | --- | --- | --- |
| A | The doctor or nurse checked that it was a convenient time to discuss research before discussing FROG | 86 (97%) | 3 (3%) | 0 (0%) |
| B | The person who spoke to me about FROG was someone who had been involved in my child's care | 58 (65%) | 15 (17%) | 16 (18%) |
| C | It was important to me that the person who spoke to me about FROG was someone involved in my child's care | 49 (55%) | 29 (33%) | 10 (11%) |
| D | The information I received about FROG was clear and straightforward to understand | 88 (99%) | 1 (1%) | 0 (0%) |
| E | I understood why my child was eligible for FROG* | 84 (96%) | 4 (5%) | 0 (0%) |
| F | I had enough opportunity to ask questions about FROG | 87 (98%) | 2 (2%) | 0 (0%) |
| G | I was satisfied with the consent process for FROG | 88 (99%) | 1 (1%) | 0 (0%) |
| H | It was difficult to take in the information I was given about FROG | 9 (10%) | 10 (11%) | 70 (79%) |
| I | It was difficult to make a decision about FROG | 7 (8%) | 20 (23%) | 62 (70%) |
| J | I made this decision | 87 (98%) | 1 (1%) | 1 (1%) |
| K | Someone took this decision away from me | 2 (2%) | 0 (0%) | 87 (98%) |
| L | I was not in control of this decision | 7 (8%) | 0 (0%) | 82 (92%) |
| M | The decision about the research was inappropriately influenced by others | 1 (1%) | 1 (1%) | 87 (98%) |

Percentages are rounded up, so some may marginally account for >100%. \*Missing data statement e) n=1

A few parents considered the information sheet too long for a stressful, time-limited situation:

> *“It was quite a lot of information. So in that moment, when you are literally making split decisions because you’re conscious that you’ve been there for a few hours, you don’t want to spend more time when your child’s agitated and unwell to read reams. So my probably feedback in that instance would be to try and create more of a one page document”* (P14, mother, interview).

Most nevertheless found the information clear, particularly when discussed with research staff: *“I think it was a good combination of having it talked through and then presenting the information”* (P6, father, interview). Only 1 parent recalled seeing a FROG poster, while 5/9 (56%) recalled the animation, which helped to *“visualise the different methods”* (P1, mother, interview).

Among 21 free-text responses from parents who declined randomisation, reasons included: concerns about distress (n=6), avoiding invasive methods (n=5) and already having obtained a CCU sample (n=4). Parents concerned about distress often favoured usual-care CCU without identifying a specific method preference: *“I do not feel comfortable putting him through this at the moment”* (P8, mother, questionnaire). A total of 3/21 specifically expressed concerns about TUBC: *“My child has had a catheter put in before and it made her extremely uncomfortable and scared”* (P77, mother, questionnaire). Conversely, 1 parent declined because they preferred TUBC: “*Normal in other countries to do catheter, I wanted the quickest option for the sample*” (P91, mother, questionnaire). Other reasons were: avoiding randomisation (n=2), a clinical decision not to proceed (n=1), uncertainty about participation (n=1) and the sample no longer being required (n=1).

#### Acceptability of urine-collection methods

Parents recruited through FROG and social media considered CCU acceptable for inclusion in a future trial. However, parents and healthcare professionals described prolonged attempts to *“wait for the clean catch”* (HCP27, research nurse, focus group 5), unsuccessful collection because CCU *“wasn’t successful”* (P16, mother, interview), and associated distress: *“She started to get quite aggravated, like, agitated at the end”* (P53, father, interview). Practical difficulties included collecting samples from a mobile child *“that just wants to move around”* (P16, mother, interview), the process being messy and ending with *“wee all over you”* (P2, mother, interview), and delays in obtaining *“the right results straightaway”* (P54, mother interview).

Most parents also considered TUBC acceptable, although some felt it was more acceptable when a child was severely unwell or old enough to understand the procedure: *“it might be easier for them to understand why a different method is going to be performed, and why they need to stay still, or all of those things, that it’s not going to hurt, this and that. It’s easier to explain”* (P33, mother, interview). Others emphasised that acceptability depended on parents understanding their child’s likely tolerance:

> *“You’ve got to know that they will tolerate it and that you’re happy to work with the timescale that it might involve as well, and have that resilience in yourself as well. You may see your child struggling but, you know, it’s under the care of a well-trained medical practitioner.”* (P54, mother, interview)

SPA was difficult to implement with most parents voicing concerns about *“the idea of a needle”* (P16, mother, interview), which was seen as unnecessary and *“really intrusive”* (P36, mother, interview) as a method of urine sampling. No interviewed parents recalled being offered SPA as a randomisation option: *“That (SPA) was never spoken about”* (P53, father, interview). Some recalled being told that it formed part of the wider study but was unavailable at their hospital:

> *“The third option (SPA) they said they weren’t able to include it in the study. I don’t know if it was just because they didn’t do it at [hospital] or what, but it wasn’t an option. So it was either the clean catch or the catheter”* (P16, mother, interview)

Most healthcare professionals confirmed this, although some reported offering SPA when clinically available:

> *“There were some instances where we did offer SPA if the clinician was happy, but a lot of the parents didn’t feel comfortable with the SPA option”* (HCP19, FG3).

Our findings were considered against the adapted Theoretical Framework of Acceptability (TFA) for paediatric trials, which consists of eight component constructs (see Table 4) [24]. The constructs of self-efficacy, trust and perceived effectiveness were fully met, whilst constructs such as affective attitude and ethicality could be fully met if SPA was not included as intervention in a future trial.

**Table 4.** Parent and healthcare practitioner acceptability of FROG mapped to the adapted theoretical framework of acceptability for paediatric trials. [**24**].

| Construct | Parent and Healthcare Practitioners Perspectives |
| --- | --- |
| **Affective attitude:** *How an individual feels about the intervention.* | Overall parents and practitioner were supportive of CCU and TUBC interventions.<br>64/99 (67%) parents consented to randomisation. Reasons for declining included concerns about distress, wishing to avoid invasive methods (TUBC). |
|  | SPA not available or offered by practitioners. TUBC was more acceptable in younger children, during acute illness, when delays in obtaining a sample could affect clinical management. |
| **Burden:** *The perceived amount of effort that is required to participate in the intervention.* | Prolonged or failed attempts at CCU. Practical difficulties in obtaining a sample in mobile children.<br>A minority declined due to perceived burden of TUBC<br>62/89 (70%) did not find it difficult to make a decision about participation. |
| **Ethicality:** The extent to which the intervention has a good fit with an individual's value system | 88/89 (99%) of parents were satisfied with the consent process.<br>Parents and practitioners viewed CCU and TUBC as appropriate and acceptable interventions in this population. SPA was not a good fit with participant value systems and described as “unnecessary” (P14, mother, interview) due to the use of such an invasive intervention for purpose of urine sampling. |
| **Intervention coherence:** The extent to which the participant understands the intervention and how it works. | 88/ 89 (99%) agreed that the information received about FROG was clear and straightforward to understand.<br>Parents had good recall and understanding of all three interventions. A few parents found the information sheet long and difficult to process when their child was unwell and agitated. Posters were rarely noticed but the animation was viewed as a helpful means of visualising the different interventions. |
| **Opportunity costs:** The extent to which benefits, profits, or values must be given up to engage in the intervention. | CCU collection was sometimes time consuming and not always successful, which either delayed or prevented access to results to inform clinical management. |
| **Perceived effectiveness:** The extent to which the intervention is perceived likely to achieve its purpose. | Parents consented for several reasons, including helping their child or other children, valuing research and trusting the practitioner introducing the study.<br>Those randomised to TUBC hoped that the intervention would speed up the process and improve the likelihood of a successful sample. |
| **Self-efficacy:** The participant's confidence that they can perform the behaviour(s) required to participate in the intervention. | 87/89 (98%) felt that they made the decision for their child to take part in the pilot trial.<br>82/89 (92%) felt in control of the decision to take part.<br>Practitioners engaged with the protocol and intervention delivery. The mean recruitment rate was 3.7 participants per site per month, exceeding the target of 3, and there were 0 losses to follow-up. |
| **Trust:** *The extent to which the participant (or parent/guardian) trusts those delivering the intervention to put the needs of patient before the requirements of the study* | Trust in practitioners delivering the study was cited by parents as a reason for providing consent for their child's participation. |

### Outcome measures

Parents and healthcare professionals identified several common priorities, including diagnostic accuracy, successful urine collection, patient discomfort or distress, adverse events and antimicrobial prescribing. However, their rankings differed.

Healthcare professionals ranked contamination as the most important primary outcome, followed by diagnostic accuracy and successful urine collection. Parents jointly ranked diagnostic accuracy and patient discomfort/distress first, followed by successful urine collection, with collection time and adverse events jointly ranked third.

For secondary outcomes, healthcare professionals prioritised collection time, discomfort/distress, parent or healthcare professional satisfaction, toleration of the sampling method, adverse events and antimicrobial prescribing. Parents identified diagnostic accuracy and discomfort/distress, followed jointly by successful collection, contamination, antimicrobial prescribing and adverse events. Satisfaction and toleration appeared only in the healthcare professionals’ secondary-outcome priorities.

### Consensus meeting

A total of 19 stakeholders attended, including 13 healthcare professionals from 8 UK hospitals and 6 parents. Consensus was defined prospectively as ≥70% agreement.

#### Population

Consensus was reached that the definitive trial should recruit children aged <18 months presenting to an emergency department with suspected UTI who were unable to provide a midstream urine sample (94% agreed; 6% disagreed).

#### Intervention and comparator

All participants agreed that the definitive trial should compare CCU with TUBC and should not include SPA (100% agreed).

#### Outcomes

Accuracy was preferred primary outcome, followed by time to sample, pain/distress, successful urine collection, appropriate antibiotic prescribing and satisfaction.

## Discussion

FROG demonstrated that a definitive trial comparing TUBC with CCU is feasible. The study exceeded its prespecified feasibility threshold (33%), with 64/170 (37.6%) eligible families consenting to randomisation. The mean recruitment rate was 3.7 participants per site per month, exceeding the target of 3, and there were 0 losses to follow-up. The perspectives study and consensus meeting further identified the population, interventions and outcomes most appropriate for a definitive trial.

Both CCU and TUBC were acceptable to parents and healthcare professionals, although each presented different challenges. CCU avoided an invasive procedure but could be slow, unsuccessful and difficult in mobile children. TUBC provided a more immediate method of collection but was associated with greater pain and distress. Its acceptability increased when children were more acutely unwell and timely sampling was considered clinically important. SPA was frequently unavailable, no families consented to randomisation involving it, and both parents and clinicians expressed concerns about its acceptability. These findings support restricting a definitive trial to CCU and TUBC, consistent with the unanimous consensus-meeting vote.

Delivery of the allocated methods was feasible but imperfect. Overall, 46/64 (71.9%) participants received their allocated method, with identical adherence in both groups. CCU was most commonly not carried out because collection times were long. TUBC was not delivered when urine could not be obtained, the child passed urine before the procedure or the procedure was abandoned because of distress. These findings are important for the design of a pragmatic definitive trial. Successful delivery will require efficient screening and recruitment processes, clear crossover criteria, staff training and timely access to clinicians trained to perform TUBC in infants and young children.

The results defined the population and setting in which a definitive comparison would be most relevant. Randomised participants were young, with a median age of 275 days; 49/64 (76.6%) were classified as higher risk and 61/64 (95.3%) were recruited from emergency departments. The perspectives findings indicated that invasive sampling was more acceptable during acute illness, when delays in obtaining a sample could affect clinical management. Consensus was therefore clear: 94% supported recruiting children aged <18 months presenting to an emergency department with suspected UTI who were unable to provide a midstream sample. This population represents the group in whom maximising the benefit of rapid, reliable sampling and minimising or avoiding any harm of an invasive procedure is most clinically relevant.

Whilst distress and pain in acutely febrile infants may be an unavoidable consequence of invasive procedures, findings from parent interviews highlighted the importance of person-, child-, and family-centred care. Consideration of the parent-child dyad, support for co-regulation, minimisation of procedural distress where possible, and maintaining resilience were identified as being of fundamental systemic importance [25].

Accuracy, speed and pain or distress emerged as the key outcomes. Healthcare professionals prioritised contamination and diagnostic accuracy, while parents placed joint emphasis on accuracy and discomfort or distress. In the consensus exercise, accuracy received the highest score, followed by time to collection and pain or distress. FROG demonstrated that contamination, collection time, pain and distress could be measured.

FROG highlighted the distinction between contamination among available cultures and its effect across the whole sampling strategy. Contamination occurred in 2/12 (16.7%) available CCU cultures and 0/6 TUBC cultures, while 2/32 (6.3%) participants allocated to CCU and 0/32 allocated to TUBC generated an observed contaminated result. The lower strategy-level rate reflects usual practice, in which not every urine sample is submitted for culture. A definitive trial should therefore prioritise clinically consequential contamination, defined as a contaminated culture prompting recall or clinical review, repeat sampling or antibiotic prescribing. This links diagnostic accuracy to outcomes experienced by children and families, while successful first-line collection, collection time, pain and distress can be assessed as key secondary outcomes.

The health-economic analysis showed that hospital resource use could be collected using case report forms and clinical records. By 30 days, 91% of the 6-month costs in the CCU group and 81% in the TUBC group had accrued. Resource use after 30 days was uncommon, although a small number of readmissions contributed to later costs. A 30-day follow-up period is therefore likely to capture most hospital costs while reducing data-collection burden, although the potential omission of infrequent later high-cost events should be considered. Only 16 ModRUM questionnaires were returned, suggesting that parent-completed questionnaires are unlikely to provide a feasible method of collecting longer-term resource use without substantial modification.

Strengths of FROG include its pragmatic multicentre design and integration of feasibility outcomes, health economics, parent and healthcare professional perspectives, and stakeholder consensus. Including families who declined randomisation provided important information about barriers to participation.

The study was not designed or powered to compare the clinical effectiveness of CCU and TUBC, and the small outcome-specific denominators preclude conclusions about differences between the methods. Many potentially eligible children were not approached because research staff were unavailable, indicating that adequate research coverage will be important in a definitive trial. SPA could not be evaluated because it was rarely available and no families consented to randomisation involving it. Parent interview participants were predominantly mothers and White British, and no children were interviewed, which may limit the perspectives represented. The health-economic analysis included randomised and non-randomised participants grouped by the method attempted and should therefore be interpreted as an assessment of data-collection feasibility rather than a randomised cost comparison.

Overall, FROG supports the feasibility of a definitive trial comparing CCU-first versus TUBC-first in children aged under 18 months presenting to emergency care with suspected UTI. The trial should be large enough to determine whether TUBC-first reduces clinically consequential contaminated cultures under usual NHS practice, while assessing successful first-line collection, collection time, pain, distress, repeat sampling and antibiotic use as secondary outcomes. Hospital resource use should be collected for 30 days.

## Sponsor

Queen’s University Belfast

## Funder

This study was funded by the National Institute for Health and Care Research Health Technology Assessment Programme (NIHR156005), awarded to Dr Thomas Waterfield. The funder had no role in the study design, data collection, analysis, interpretation or preparation of the manuscript.

## Trial Management

Northern Ireland Clinical Trials Unit

## Trial Registration

ISRCTN84676764, registered on 1 April 2025

## Ethical Approval

North East – Newcastle & North Tyneside 1 Research Ethics Committee, Ref: 24/NE/0222

Additional files related to this study are available at: ISRCTN84676764.

## Data availability

The participant-level dataset has not been deposited in a public repository because the small sample size and detailed clinical information create a risk of participant re-identification. Data with direct identifiers removed may be made available upon reasonable request to the corresponding author. Requests should describe the proposed use and data required and will be considered in accordance with participant consent, ethical and data-protection requirements, sponsor approval and completion of an appropriate data-sharing agreement

## Supporting information

COREQ Checklist

CONSORT Checklist

## References

1. Becknell B, Schober M, Korbel L, et al. The diagnosis, evaluation and treatment of acute and recurrent pediatric urinary tract infections. Expert Rev Anti Infect Ther. 2015;13(1):81–90.

2. Coulthard MG, Lambert HJ, Keir MJ. Occurrence of renal scars in children after their first referral for urinary tract infection. BMJ. 1997;315(7113):918–919.

3. National Institute for Health and Care Excellence. Urinary tract infection in under 16s: diagnosis and management. NICE guideline [NG224]. Published 27 July 2022. Available at: https://www.nice.org.uk/guidance/ng224. Accessed 21 August 2026.

4. Pantell RH, Roberts KB, Adams WG, et al. Evaluation and management of well-appearing febrile infants 8 to 60 days old. Pediatrics. 2021;148(2):e2021052228.

5. Shaikh N, Craig JC, Rovers MM, et al. Identification of children and adolescents at risk for renal scarring after a first urinary tract infection: a meta-analysis with individual patient data. JAMA Pediatr. 2014;168(10):893–900.

6. Velasco R, Lejarzegi A, Gomez B, et al. Febrile young infants with abnormal urine dipstick at low risk of invasive bacterial infection. Arch Dis Child. 2021;106:758–763.

7. Gomez B, Mintegi S, Bressan S, et al. Validation of the “Step-by-Step” approach in the management of young febrile infants. Pediatrics. 2016;138(2):e20154381.

8. Buettcher M, Trueck J, Niederer-Loher A, et al. Swiss consensus recommendations on urinary tract infections in children. Eur J Pediatr. 2021;180(3):663–674.

9. Robinson JL, Finlay JC, Lang ME, et al. Urinary tract infections in infants and children: diagnosis and management. Paediatr Child Health. 2014;19(6):315–325.

10. Tosif S, Baker A, Oakley E, et al. Contamination rates of different urine collection methods for the diagnosis of urinary tract infections in young children: an observational cohort study. J Paediatr Child Health. 2012;48:659–664.

11. Branagan A, Canty N, O’Halloran E, et al. Evaluation of the Quick-Wee method of inducing faster clean catch urine collection in pre-continent infants: a randomised controlled trial. World J Pediatr. 2022;18(1):43–49.

12. Kaufman J, Fitzpatrick P, Tosif S, et al. Faster clean catch urine collection (Quick-Wee method) from infants: randomised controlled trial. BMJ. 2017;357:j1341.

13. Tzimenatos L, Mahajan P, Dayan PS, et al. Accuracy of the urinalysis for urinary tract infections in febrile infants 60 days and younger. Pediatrics. 2018;141:e20173068.

14. World Health Organization. Antimicrobial resistance. Available at: https://www.who.int/health-topics/antimicrobial-resistance. Accessed 21 August 2026.

15. World Health Organization. Antimicrobial resistance: global report on surveillance. Geneva: World Health Organization; 2021. Available at: https://www.who.int/publications/i/item/9789240027336. Accessed 21 August 2026.

16. Taylor Miller P, McDowell C, Agus A, et al. Determining the feasibility of randomising infants, children and young people to invasive and non-invasive urine sampling techniques (FROG): protocol for a multicentre randomised controlled feasibility trial and mixed methods perspectives’ study of RCT feasibility. NIHR Open Res. 2025;5:108. doi:10.3310/nihropenres.14114.1.

17. Eldridge SM, Chan CL, Campbell MJ, Bond CM, Hopewell S, Thabane L, Lancaster GA, on behalf of the PAFS consensus group. CONSORT 2010 statement: extension to randomised pilot and feasibility trials. BMJ. 2016;355:i5239.

18. Garfield KM, Thorn JC, Noble SM, Husbands SK, Hollingworth W. Development of a brief, generic, modular resource-use measure (ModRUM): piloting with patients. BMC Health Serv Res. 2023;23:994.

19. Husereau D, Drummond M, Augustovski F, et al. Consolidated Health Economic Evaluation Reporting Standards 2022 (CHEERS 2022) statement: updated reporting guidance for health economic evaluations. Value Health. 2022;25(1):3–9.

20. Mitchell TK, Menzies JC, Ramnarayan P, et al. Developing an adaptive paediatric intensive care unit platform trial with key stakeholders: a qualitative study. BMJ Open. 2025;15(1):e085142. doi:10.1136/bmjopen-2024-085142.

21. Lansdale N, Woolfall K, Deja E, et al. Timing of Stoma Closure in Neonates: the ToSCiN mixed-methods study. Health Technol Assess. 2024;28(71):1–130. doi:10.3310/JFBC1893.

22. Malterud K, Siersma VD, Guassora AD. Sample size in qualitative interview studies: guided by information power. Qual Health Res. 2016;26(13):1753–1760. doi:10.1177/1049732315617444.

23. Byrne D. A worked example of Braun and Clarke’s approach to reflexive thematic analysis. Qual Quant. 2022;56(3):1391–1412. doi:10.1007/s11135-021-01182-y.

24. Deja E, Peters MJ, Khan I, et al. Establishing and augmenting views on the acceptability of a paediatric critical care randomised controlled trial (the FEVER trial): a mixed methods study. BMJ Open. 2021;11(3):e041952. doi:10.1136/bmjopen-2020-041952.

25. Segers, E.W., van den Hoogen, A., Schoonhoven, L. et al. How to meet coping strategies and preferences of children during invasive medical procedures: perspectives of healthcare professionals. Eur J Pediatr. 2024;183:5291–5301. doi:10.1007/s00431-024-05802-1.

